# Predicting maternal social loneliness by passive sensing with wearable devices

**DOI:** 10.1101/2022.10.10.22280910

**Authors:** Fatemeh Sarhaddi, Iman Azimi, Hannakaisa Niela-Vilén, Anna Axelin, Pasi Liljeberg, Amir M. Rahmani

## Abstract

**Background:** Maternal loneliness is associated with adverse physical and mental health outcomes for both the mother and her child. Detecting maternal loneliness non-invasively through wearable devices and passive sensing provides opportunities to prevent or reduce the impact of loneliness on the health and well-being of the mother and her child.

**Objective:** The aim of this study is to use objective health data collected passively by a wearable device to predict maternal (social) loneliness during pregnancy and the postpartum period based on and to identify the important objective physiological parameters in loneliness detection.

**Methods:** We conducted a longitudinal study using smartwatches to continuously collect physiological data from 31 women during pregnancy and the postpartum period. The participants completed the University of California, Los Angeles (UCLA) loneliness questionnaire in gestational week 36 and again at 12 weeks postpartum. Responses to this questionnaire and the background information of the participants were collected via our customized cross-platform mobile application. We leveraged participants’ smartwatch data from the 7 days before and the day of their completion of the UCLA questionnaire for loneliness prediction. We categorized the loneliness scores from the UCLA questionnaire as loneliness (scores ≥ 12) and non-loneliness (scores *<* 12). We developed decision tree and gradient boosting models to predict loneliness. We evaluated the models by using a leave-one-participant-out cross validation. Moreover, we discussed the importance of extracted health parameters in our models for loneliness prediction.

**Results:** The gradient boosting and decision tree models predicted maternal social loneliness with weighted F1 scores of 0.871 and 0.897, respectively. Our results also show that loneliness is highly associated with activity intensity, activity distribution during the day, resting heart rate (HR), and resting heart rate variability (HRV).

**Conclusion:** Our results show the potential benefit and feasibility of using passive sensing with a smartwatch to predict maternal loneliness. Our developed machine learning models achieved a high F1 score for loneliness prediction. We also show that intensity of activity, activity pattern, and resting HR and HRV are good predictors of loneliness. These results indicate the intervention opportunities made available by wearable devices and predictive models to improve maternal well-being by early detection of loneliness.

## Introduction

Loneliness is a subjective unpleasant feeling of mismatch between desired and perceived meaningful social relationships [1]. This feeling consists of social and emotional dimensions: social loneliness refers to a lack of an engaging social network or a desired group of contacts, whereas emotional loneliness is caused by a lack of intimate relationships or close emotional attachments [2]. Loneliness can have adversarial health consequences such as negative cardiovascular outcomes and mental health disorders [3] and even increases the risk of mortality [4]. In addition, loneliness is a global public health issue that is growing in modern society and has increased especially during the COVID-19 pandemic and its attendant social isolation [5].

Maternal loneliness during pregnancy and the postpartum period is associated with several health issues for the mother and her child. Various studies showed a positive correlation between loneliness and depression during pregnancy across countries [5–10]. In addition, maternal loneliness is associated with life dissatisfaction and pair-relationship dissatisfaction [9]. Other studies showed that loneliness is significantly associated with postpartum depression [11, 12]. It was also shown that, in the COVID-19 pandemic, loneliness during pregnancy was associated with serious psychological distress [13], anxiety [14], cognitive distortion [10], higher level of perceived stress [6], and lower level of social support [6, 7]. Maternal loneliness increases the risk of respiratory tract infections in newborn babies [15]. The prediction or early detection of maternal loneliness could help avoid adversarial consequences for the mother and her child through proper intervention.

Previous studies investigated loneliness during pregnancy and the postpartum period utilizing observational methods based on self-report measures, such as standard questionnaires [8] and interviews [16]. For example, Perzow *et al*. in [5] used self-report questionnaires to discern symptoms of depression and anxiety, loneliness, and COVID-19-related adverse health outcomes. In another study, Giurgescu *et al*. [6] used online surveys to investigate the association between loneliness, depression, perceived stress, and social support during the COVID-19 pandemic in pregnant Black women. In [10], standard questionnaires were utilized to study the relationship among loneliness, depression, and cognitive distortion. These studies investigated the associations between loneliness and various health issues, comparing the loneliness of people with health problems to the loneliness of those without health problems. However, they do not recommend or describe proactive services to predict or detect loneliness early on. In addition, subjective studies require participants’ engagement in answering the questionnaires or interview questions. Therefore, data collection is burdensome for pregnant women, especially in late pregnancy or during the postpartum period when they are occupied with a newborn baby and may find it difficult to remember and find time to answer questionnaires or engage in an interview.

Using wearable devices and smartphones for well-being and healthcare applications has been increasing rapidly in recent years. These devices enable continuous passive sensing of socio-behavioral data. However, few studies have utilized smartphones and wearable devices to *predict* using passive sensing [17, 18]. The authors of one study [18] leveraged GPS and Bluetooth data, gathered by participants’ smartphones, to explore the association between momentary loneliness and companionship type in college students. In addition, another study [17] explored the sleep and physical activity data recorded on a wristband activity tracker, as well as the location, screen time, calls and SMS logs, and Bluetooth data of college students, over the course of a semester and used this data to predict loneliness. Although these studies predicted loneliness by using wearable devices and passive sensing, they were limited to college students living on a university campus. Moreover, these studies did not use heart rate variability (HRV) features, even though it has been shown that loneliness is associated with lower resting HRV [19].

To the best of our knowledge, there is no study in the literature that has predicted maternal loneliness during pregnancy and the postpartum period on the basis of objective physiological data. The previously mentioned studies were limited to subjective data or performed on other population groups (i.e., college students). Predicting maternal loneliness with the use of passive data sensing is beneficial to improving maternal and child well-being with minimal cost and effort required of mothers.

In this paper, we present a passive sensing method, enabled by a smartwatch, for loneliness prediction during late pregnancy and the postpartum period. The smartwatch collected heart rate (HR), HRV, physical activity, and sleep parameters. These physiological parameters were chosen due to the association of loneliness with lower resting HRV [19], decreased physical activity [20], and poor self-reported sleep quality [21, 22]. We then developed two machine learning models - decision tree and gradient boosting - to predict loneliness based on the objective data. Moreover, we investigated the importance of health parameters in loneliness prediction. In summary, the main contributions of this paper are as follows:

- Presenting a passive sensing method, enabled by a smartwatch, for loneliness prediction during late pregnancy and the postpartum period
- Developing two machine learning models to predict loneliness during pregnancy and the postpartum period based on objective health data collected by a wearable device
- Investigating and discussing physiological parameters’ importance to maternal loneliness prediction.

## Materials and methods

### Study design

An observational longitudinal study was conducted in free-living conditions with a convenience sample of pregnant women in Southwest Finland. This study is part of a project utilizing a wearable-based system to remotely monitor women’s physiological health parameters, including HR, HRV, sleep, and physical activity during pregnancy and the postpartum period. This system used a smartwatch to collect objective health parameters and a cross-platform mobile application to collect subjective and background information. The remote maternal monitoring system was described and evaluated in a previous publication [23].

### Participants and recruitment

Pregnant women with singleton pregnancies were recruited at 12–15 gestational weeks for this study. The inclusion criteria were: 1) having the ability to understand the Finnish language, 2) being at least 18 years old, and 3) having an Android or iOS smartphone.

Recruitment was performed via maternity clinics or social media advertisements for two groups of pregnant women with different inclusion criteria from January 2019 to March 2020. The first group included women with a history of preterm birth (gestational weeks 22–36) or late miscarriage (gestational weeks 12–22). The second group consisted of women with a history of previous full-term uncomplicated pregnancy and no pregnancy losses.

In scheduled face-to-face meetings with eligible volunteer pregnant women, the researchers informed the women about the study. Then, participants provided their written informed consent and received a smartwatch and the study instructions. The participants were asked to wear the smartwatch continuously during their pregnancies and for three months postpartum. They also installed our customized cross-platform mobile application on their smartphones. Sixty-two pregnant women were recruited for this study. Four women withdrew from the study. We also excluded data from participants with a high amount of missing data (see see the section titled Datasets and machine learning models for loneliness prediction). Thus, 31 pregnant women were included in this study. The participants’ background information is provided in Table 1.

**Table 1.** Participant background information (n = 31)

| Parameter | Value |
| --- | --- |
| Age mean (SD) | 31.9 (4.9) |
| BMI mean (SD) | 25.98 (5.96) |
| Marital status |  |
| Married or cohabitation | 98.3 % |
| Other | 1.7 % |
| Work status |  |
| Working | 75.9 % |
| Student | 12.1 % |
| Unemployed | 1.7% |
| Other | 10.3 % |
| Education |  |
| High school | 41.3 % |
| College | 31.1 % |
| University | 27.6 % |
| Pregnancy weeks at birth, mean (SD) | 36+4 (9+6) |

### Research ethics

This study received ethical approval from the Ethics Committee of the Hospital District of Southwest Finland, approval number: Dnro: 1/1801/2018. Written informed consent was obtained from all participants.

### Data Collection

Data were collected from each participant by a Samsung Gear Sport smartwatch and by our customized cross-platform mobile application. The Samsung smartwatch included a photoplethysmography (PPG) sensor and an inertial measurement unit. It ran Tizen OS, which is an open-source operating system that enabled us to develop customized applications for the smartwatch. The smartwatch provided PPG signals, acceleration data, and gyroscope data. We developed customized applications for the smartwatch to collect sleep and physical activity data continuously and collect 12 minutes of PPG signal every other hour. The data were stored on the internal storage of the smartwatch. In addition, we developed a smartwatch application to transfer the collected data to our cloud server via Wi-Fi. The smartwatch, enabled by our applications, had sufficient battery life (i.e., 2–3 days) for data collection [23].

Our customized cross-platform mobile application provided self-report questionnaires to the participants. To evaluate loneliness, we used the 12-item version of the Revised UCLA Loneliness Scale questionnaire consisting of questions about the factors of social and emotional loneliness [24]. Each factor was addressed by 6 questions to which the answers had potential scores from 6 to 24. Higher points indicated greater feelings of loneliness. The participants completed structured questionnaires at two time points: at gestational week 36 and at three months postpartum. We also collected background information through the mobile application.

### Datasets and machine learning models for loneliness prediction

The collected data from the smartwatch were used to generate seven datasets. Then, we developed two machine learning models and utilized these datasets to train and test our models. Finally, we investigated the important parameters for loneliness prediction. Our machine learning pipeline, of which an overview is shown in Fig 1, comprised the following process:

**Fig 1.**
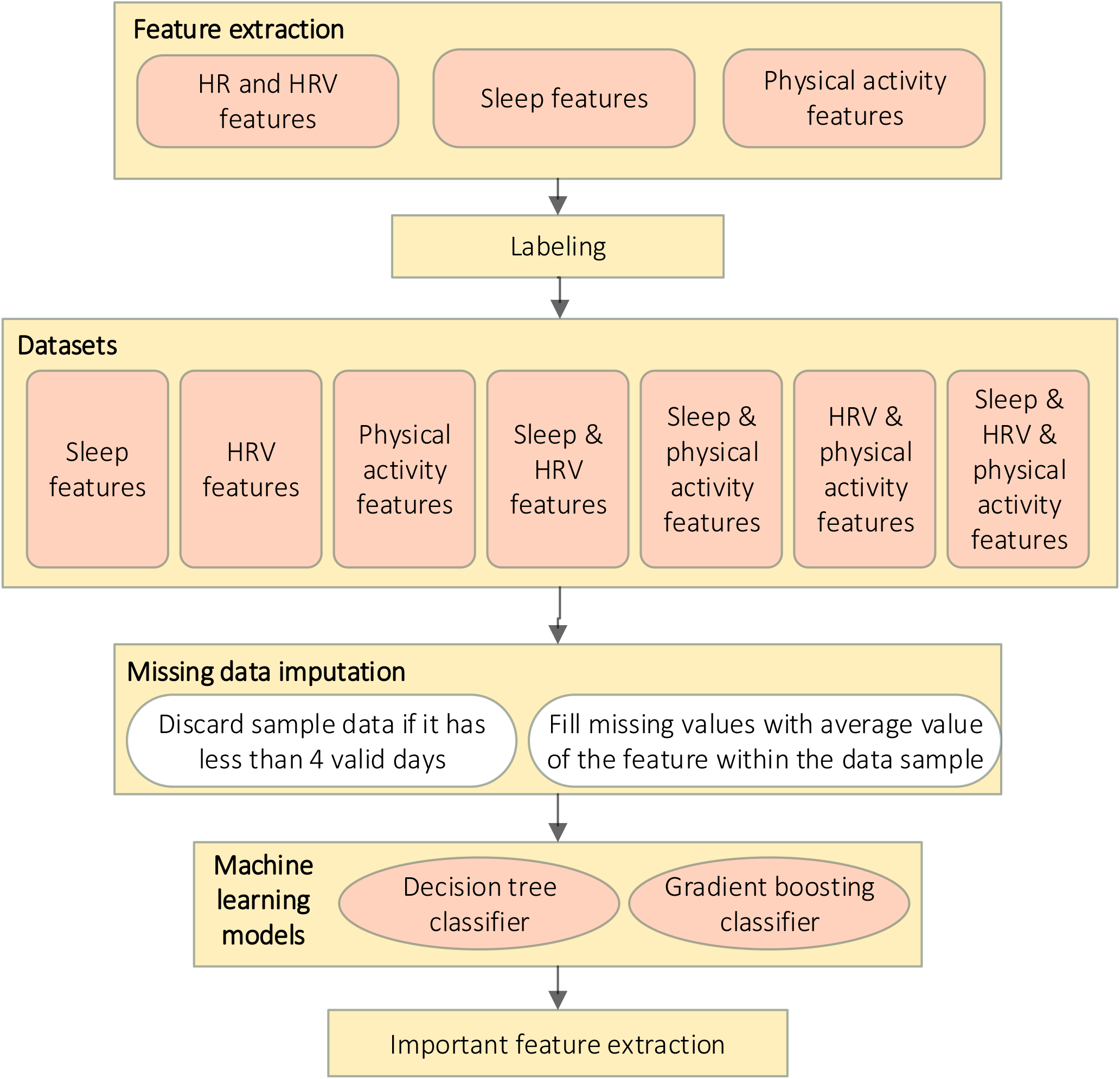
Machine learning pipeline

1. Feature extraction
2. Dataset creation and labeling
3. Missing data imputation
4. Training and testing the machine learning models (decision tree and gradient boosting) for different datasets
5. Investigating the important features of the two machine learning models for loneliness prediction

These steps are described in the following sections.

#### Feature extraction

We extracted HR and HRV, sleep, and physical activity data from the objective data collected by the smartwatch.

#### HR and HRV features

We utilized PPG signals to extract HR and HRV features. The smartwatch collected 12 minutes of PPG signals every other hour with a sampling frequency of 20 Hz (as described in a previous publication [23]). Based on the duration of PPG recordings, we used short-term HRV analysis, including 5-minute windows of PPG signals for HRV extraction [25, 26]. Our HR and HRV extraction pipeline consisted of three steps:

1. **Reliable signals detection:** PPG signals are prone to noise, such as motion artifacts. Therefore, unreliable signals had to be detected and discarded. We trained a support vector machine (SVM) classifier to distinguish reliable and unreliable signals. The SVM classifier was trained using several morphological features of the PPG signals, including skewness, kurtosis, approximate entropy, Shannon entropy, and spectral entropy [27]. Then, we leveraged the trained model to detect and subsequently discard unreliable PPG signals.
2. **Peak detection and interbeat interval (IBI) extraction:** We used a bandpass filter with cut-off frequencies of 0.7 Hz and 3.5 Hz to filter out noises outside the human heart rate range. We used a moving average-based peak detection method with adaptive thresholds to detect peaks and extract IBIs. Then, we utilized error detection methods to remove false peaks and their corresponding IBIs. To this end, too-large or too-small IBIs were removed based on the other IBIs in the same window of the signals. The peak detection and IBI extraction method was implemented using HeartPy library [28] in Python.
3. **Resting HR and HRV extraction:** We extracted resting HR (when the HR is the lowest during sleep) and its corresponding HRV parameters using detected peaks and extracted IBIs.HR is calculated as the number of peaks per minute. We utilized normal IBIs to extract HRV parameters that could be reliably extracted at the sampling frequency of collected PPG signals (20 Hz) [29]. The extracted HRV parameters were average normal IBIs (AVNN), root mean square of the successive differences (RMSSD), standard deviation of IBIs (SDNN), power in low-frequency range (LF), power in high-frequency range (HF), and LF to HF ratio (LF/HF).

#### Sleep features

Using the smartwatch, we recorded total sleep time (TST), sleep fragmentation, wake after sleep onset (WASO), and average hand movement during sleep, as described in a previous publication [30]. We also added a sleep quality indicator showing WASO ≤ 20 minutes and a sufficient sleep parameter, which showed TST between 7 and 8.5 hours.

#### Physical activity features

The smartwatch captured several physical activity parameters at a granularity of 10 minutes. By aggregating the smartwatch’s activity parameters during participants’ awake time, we extracted daily step counts, walking steps, running steps, distance, activity duration, and activity intensity. We then calculated the sedentary time as awake time without walking and running activities. We also added a sufficient activity indicator to show daily step counts above 7000. Finally, we extracted statistical parameters from the distribution of step counts and duration of activity during the day, based on the hourly data. The statistical parameters of hourly activity distribution were mean, minimum, median, maximum, standard deviation (SD), interquartile range, range, skewness, kurtosis, and root mean square.

Table 2 shows the summary of extracted features.

**Table 2.**
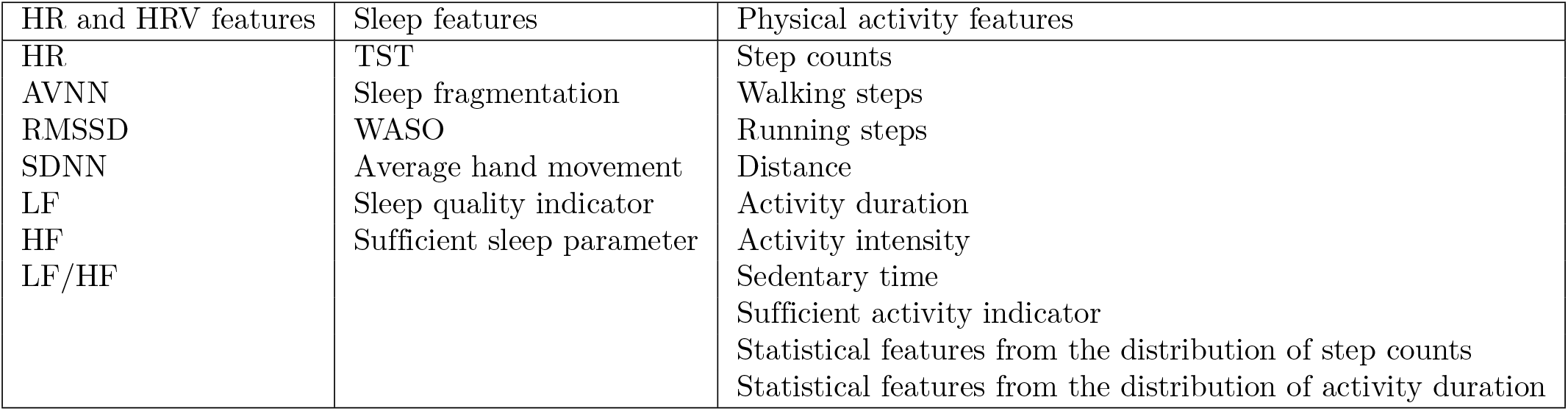
Summary of extracted features

## Dataset creation and labeling

We generated seven datasets using different combinations of HR and HRV, sleep, and physical activity features extracted from the smartwatch data. These datasets were used to train and test our developed machine learning models.

Labeling was performed utilizing UCLA social scores. We used binary classification for loneliness prediction and considered UCLA social score ≥ 12 as 1 (loneliness) and UCLA social score *<* 12 as 0 (no loneliness) [24]. We ignored the UCLA emotional scores since we had few participants with emotional loneliness. We used UCLA participants’ responses at gestational week 36 and at week 12 after delivery.

Each sample in our datasets contained data from 7 days before and on the day of answering the UCLA questionnaires as the UCLA questions ask respondents to consider their feelings over the previous week.

### Missing data imputation

We added a data sample which contained 8 days of data and a loneliness label from one participant in the datasets if the data sample had at least 4 valid days. A *valid day* was defined as a day in which the participant wore the smartwatch for at least 10 hours during waking hours and in which the watch collected valid sleep data. The samples with fewer than 4 valid days were discarded due to the high proportion of missing data. In addition, we used the average values of each feature in one data sample to fill the missing values in that data sample.

We added 39 data samples from 31 participants (eight participants had two data samples) to our datasets. The data from other participants were excluded due to the high ratio of missing data. These data were missing as a result of different technical and practical issues during monitoring. For example, some participants had preterm births before gestational week 36. Many wore the watch for an insufficient amount of time. Some participants removed the smartwatch’s customized application by resetting the watch.

### Machine learning models

We developed decision tree and gradient boosting models for predicting loneliness and investigating the importance of features for loneliness prediction.

A decision tree classifier is a simple, flexible, robust, and easy to interpret method, which is well suited to complex ecological data [31, 32]. This model has a tree-like structure which includes internal nodes and leaves. Each internal node splits the data based on one feature. The features are selected based on the Gini index, which represents the purity of classification. Each leaf node shows the class label. The decision tree method was chosen as it is fast and simple, and a specific feature’s importance can be easily understood from the tree structure.

Gradient boosting is another machine learning method that can be used for classification and regression. It is an ensemble of weak prediction models, and in each step of training, it adds a new estimator to improve the results. This model performs well on noisy data and outperforms most common machine learning models [33]. This model has performed well in prediction with nonlinear decision boundaries and has produced good results in similar studies [17].

#### Model evaluations

We investigated the performance of the predictive machine learning models using the leave-one-participant-out cross-validation method and reported the average performance thereof. Using the scikit-learn library in Python [34], we developed and evaluated the following aspects of the models:

The following machine learning measures were used for performance evaluation:

- Precision: percentage of predicted samples that actually belonged to a class
- Recall: percentage of correctly predicted samples per class
- F1 score: harmonic mean of precision and recall per class
- Weighted F1 score: weighted average of F1 scores

## Results

In this section, we present the performance of our predictive models in terms of precision, recall, F1 score, and weighted F1 scores. We also discuss the importance of features in loneliness prediction.

### Loneliness prediction

The prediction results of the predictive models for different datasets are summarized in Table 3. The datasets contain HRV features, sleep features, physical activity features and different combinations of these feature sets.

**Table 3.** Per class precision, recall and F1 score, and weighted F1 score performance measures for the predictive models

| Model | Dataset | Precision |  | Recall |  | F1 score |  | Weighted F1 score |
| --- | --- | --- | --- | --- | --- | --- | --- | --- |
|  |  | not lonely | loneliness | not lonely | loneliness | not lonely | loneliness |  |
| Decision Tree | Sleep | 0.632 | 0.5 | 0.585 | 0.541 | 0.585 | 0.541 | 0.566 |
|  | HRV | 0.545 | 0.459 | 0.545 | 0.459 | 0.545 | 0.459 | 0.506 |
|  | PA | **0.909** | **0.882** | **0.909** | **0.882** | **0.909** | **0.882** | **0.897** |
|  | Sleep & HRV | 0.609 | 0.5 | 0.636 | 0.471 | 0.622 | 0.485 | 0.562 |
|  | Sleep & PA | **0.905** | **0.833** | **0.864** | **0.882** | **0.884** | **0.857** | **0.872** |
|  | HRV & PA | **0.909** | **0.882** | **0.909** | **0.882** | **0.909** | **0.882** | **0.897** |
|  | ALL | **0.909** | **0.882** | **0.909** | **0.882** | **0.909** | **0.882** | **0.897** |
| Gradient Boosting | Sleep | 0.526 | 0.4 | 0.455 | 0.471 | 0.488 | 0.432 | 0.464 |
|  | HRV | 0.532 | 0.441 | 0.568 | 0.405 | 0.549 | 0.423 | 0.491 |
|  | PA | 0.792 | 0.8 | 0.864 | 0.706 | 0.826 | 0.75 | 0.793 |
|  | Sleep & HRV | 0.556 | 0.429 | 0.455 | 0.529 | 0.5 | 0.474 | 0.488 |
|  | Sleep & PA | 0.769 | 0.846 | 0.909 | 0.647 | 0.833 | 0.733 | 0.790 |
|  | HRV & PA | **0.87** | **0.875** | **0.909** | **0.824** | **0.889** | **0.848** | **0.871** |
|  | ALL | **0.833** | **0.867** | **0.909** | **0.765** | **0.87** | **0.812** | **0.845** |

The decision tree model achieved the best performance on datasets that contained only physical activity features, physical activity, and HRV features, or all the features. The results for the dataset with physical activity and sleep were slightly lower than the results for physical activity features. In addition, the decision tree model performed poorly (weighted F1 score less than 57%) on datasets of HRV features, sleep features, and the combination of HRV and sleep features. These results show that physical activity features had the highest impact on the prediction results for the decision tree model and that sleep features impacted the prediction results negatively. Fig 2 shows the decision tree model for the physical activity features dataset and the features used in the loneliness predictions.

**Fig 2.**
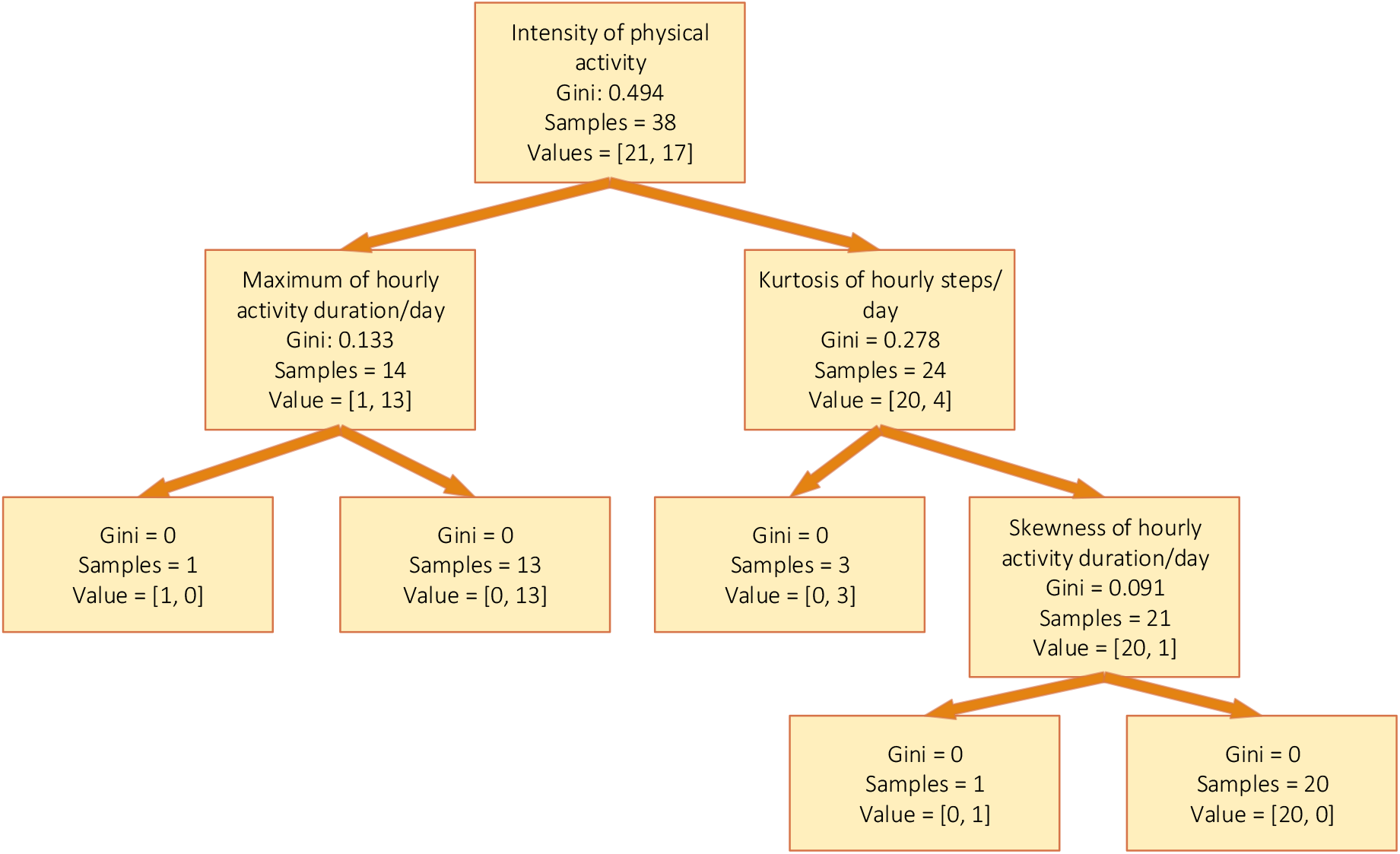
Features used in decision tree model for physical activity datasets

**Fig 3.**
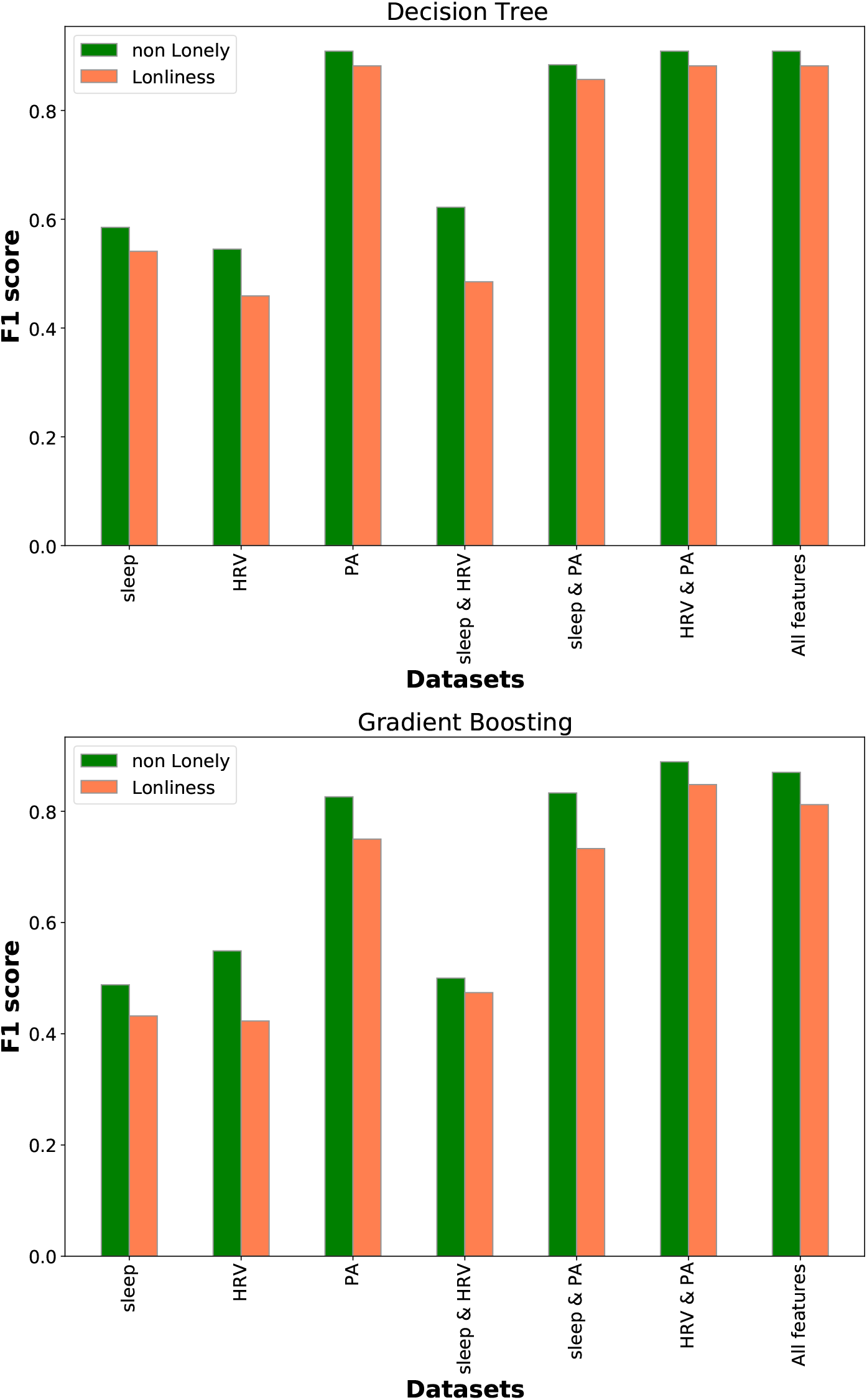
Classification results for different datasets

As did the decision tree model, the gradient boosting model had better classification results for datasets containing physical activity than for other datasets. Moreover, gradient boosting performed better on the dataset of physical activity features than it did on the dataset of physical activity and sleep features. The model also performed better on the dataset of all features compared with its performance on dataset containing only physical activity. However, the gradient boosting model achieved higher performance on the dataset containing both physical activity and HRV features than it did on any other dataset.

### Feature importance in loneliness prediction

We investigated the importance of the features in the decision tree and gradient boosting models on datasets that achieved a weighted F1 score higher than 80%. For the decision tree, in the four models with the highest F1 score, the most important features were intensity of activity and kurtosis of the steps during the day (based on hourly data). Other important features were resting SDNN, HF, RMSSD, maximum, interquartile range, and skewness of duration of activity during the day. For gradient boosting, the most important features were intensity of activity, several distribution parameters of total steps, and duration during the day, including kurtosis, maximum, average, interquartile range, SD, range, and root mean square. In addition, HRV features (including resting HR, SDNN, RMSSD, HF, and LF/HF) had a high level of importance in the gradient boosting model.

The most frequently selected features in these models show their significant impact on prediction. Therefore, the results show that intensity of activity, activity distribution during the day, and resting HR and HRV have the highest association with and effect on loneliness.

## Discussion

### Principal findings

In this study, we developed two predictive models – decision tree and gradient boosting – to predict loneliness during late pregnancy and the postpartum period by using physiological data collected by a smartwatch. The models used 8 days of data, collected passively by a smartwatch, to predict maternal social loneliness. The gradient boosting and decision tree models achieved weighted F1 scores of 0.871 and 0.897, respectively. These results show the feasibility of predicting maternal loneliness during pregnancy and the postpartum period by passive sensing using wearable devices.

In addition, we investigated the importance of sleep, resting HR and HRV, and physical activity collected by the smartwatch for loneliness prediction. Our results show that physical activity, patterns of activity during the day, and resting HR and HRV are the most important predictors of loneliness. Moreover, sleep features have no effect on prediction results. The decision tree results show that having high or intensive activity levels (i.e., when most of a participant’s daily steps happen within a short period of time) can be a good sign of non-loneliness. On the other hand, having less intensive activity levels and low resting HRV when most of a participant’s activity takes place before evening can be a predictor of loneliness.

This finding about the association between low physical activity and increased loneliness is very important for maternity care. It is well known that women’s levels of physical activity decrease as pregnancy proceeds [35]; by contrast, high levels of prenatal activity and exercise are associated with lower pregnancy-related and obstetric complications as well as with higher health-related quality of life [36–38]. Though a low level of physical activity may, in itself, be a risk for many adverse outcomes, it could be also a sign of loneliness and thereby further increase negative health consequences.

Therefore, health care professionals should encourage pregnant women to be physically active but, simultaneously, should be attentive to the signs of loneliness so that they are able to support pregnant women individually and, by implication, promote the health of both the mother and her fetus/infant.

### Comparison with previous studies

To the best of our knowledge, this is the first study predicting loneliness during pregnancy and the postpartum period based on objective health parameters. Previous work usually considered college students/young adults [17, 18, 39] and elderly people [40].

Badal *et al*. [40] used natural language processing methods to predict loneliness in elderly people. Our results show higher precision, recall, and F1 scores than their results for quantitative loneliness prediction. Moreover, their method required a semi-structured interview. However, our method passively collects data and requires no further effort from participants. In another study [18], researchers used GPS and Bluetooth data gathered by participants’ smartphones, as well as ecological momentary assessment surveys collecting real-time self-report information about companionship types and social interactions. Their models can predict self-report loneliness with an average area under the curve (AUC) equal to 0.74. In contrast, our models have better performance and we used a standard UCLA questionnaire for labeling. In another study [17], Doryab *et al*. predicted loneliness for college students with an accuracy of 80.2%, based on data collected from a smartphone and a wearable device. Our results for pregnant women achieve higher performance than their work did. Moreover, their model requires the use of more information from participants (such as the phone numbers of close friends or family members, used to assess calls to close contacts) that raises privacy concerns. However, our work only used physiological parameters.

In addition, some studies investigated important features in loneliness prediction. Wang *et al*. [39] showed that daily activity duration, traveled distance, and activity duration in the evening are negatively correlated with loneliness in college students. Other studies also showed the negative correlation between loneliness and duration of activity and total movements and step counts [17, 41]. This is in alignment with our results that show physical activity features to be the most important factors in loneliness predictions. The authors of another study [42] reported that loneliness was not associated with sleep duration, a result confirmed by our study’s finding regarding participants’ sleep parameters.

### Limitation and future work

We have 39 valid data samples with which to train and test our predictive models. In the future, we need to test our predictive models with more data in order to generalize the results. In addition, the participants in our study were healthy. Therefore, the predictive models’ validity is limited to the healthy population. In the future, we should consider including participants with diagnosed health problems.

We used data from late pregnancy (gestational week 36) and 12 weeks postpartum for loneliness prediction. However, it is known that physiological health parameters such as HRV and physical activity change during pregnancy and the postpartum period [43, 44], e.g., physical activity decreases during pregnancy. Generalizing our predictive models to the whole pregnancy requires using data from additional weeks during pregnancy and the postpartum period.

## Conclusion

In this paper, we presented predictive machine learning models for loneliness prediction during pregnancy and the postpartum period. Utilizing HRV, sleep, and physical activity data collected by smartwatches, our presented predictive models achieved high F1 scores. Our findings illustrate the potential benefit and feasibility of predicting loneliness during pregnancy by using objective data collected passively through a smartwatch. In addition, our findings provide insight into which physiological parameters are associated with loneliness during late pregnancy and the postpartum period. Using passive sensing and predictive models to predict and detect loneliness can support the creation of interventions based on prediction outcomes and thereby effectively improve maternal and infant well-being and prevent adverse health outcomes related to loneliness.

## Data Availability

The data used in this study include sensitive health information, and the informed consent signed by the participants does not allow the data to be made publicly available due to ethical restriction. According to the current approval by the Ethics Committee of the Hospital District of Southwest Finland, the participants gave permission to use the collected data only for the purpose described in the consent. Data requests may be subject to individual consent and/or ethics committee approval. Researchers wishing to use the data should contact the Ethics Committee of the Hospital District of Southwest Finland, contact details: Tyks U-hospital, Kiinamyllynkatu 4-8, UB3, PO Box 52, FI-20521 TURKU, Finland. We recommend first to contact the PI of the research project, associate professor Anna Axelin, contact details: University of Turku, Department of Nursing Science, 20014 University of Turku, Finland.

